# Quality of life in cancer patients treated with mistletoe: a systematic review and meta-analysis

**DOI:** 10.1101/19013177

**Authors:** Martin Loef, Harald Walach

## Abstract

**Background:** Mistletoe extracts are used as an adjunct therapy for cancer patients, but there is dissent as to whether this therapy has a positive impact on quality of life (QoL).

**Methods:** We conducted a systematic review searching in several databases (Medline, Embase, CENTRAL, CINAHL, PsycInfo, Science Citation Index, clinicaltrials.gov, opengrey.org) by combining terms that cover the fields of “neoplasm”, “quality of life” and “mistletoe”. We included prospective controlled trials that compared mistletoe extracts with a control in cancer patients and reported QoL or related dimensions. The quality of the studies was assessed with the Cochrane Risk of Bias tool version 2.We conducted a quantitative meta-analysis.

**Results:** We included 26 publications with 30 data sets. The studies were heterogeneous. The pooled standardized mean difference (random effects model) for global QoL after treatment with mistletoe extracts vs. control was d = 0.61 (95% CI 0.41-0.81; p<0,00001). The effect was stronger for younger patients, with longer treatment, in studies with lower risk of bias, in randomized and blinded studies. Sensitivity analyses support the validity of the finding. 50% of the QoL subdomains (e.g. pain, nausea) show a significant improvement after mistletoe treatment. Most studies have a high risk of bias or at least raise some concern.

**Conclusion:** Mistletoe extracts produce a significant, medium-sized effect on QoL in cancer. Risk of bias in the analyzed studies is likely due to the specific type of treatment, which is difficult to blind; yet this risk is unlikely to affect the outcome.

**PROSPERO registration number:** CRD42019137704

## Background

Mistletoe has been used for centuries as a folk remedy, dating back to ancient Greek and Celtic medicine [1, 2]. Today’s use of preparations of the European mistletoe (*Viscum album L*.) in anthroposophic medicine and particularly the treatment of cancer patients has been introduced by Rudolf Steiner (1861-1925) in the 1920s [3] and developed further by Ita Wegmann and various clinicians and pharmacists in this tradition. The basis for this development were, apart from the traditional pharmacopeias, specific anthroposophic investigations involving the phenomenology of mistletoe that grows as a semi-parasite on trees. These and other considerations led Steiner and associates to try mistletoe treatment for cancer. Out of this suggestion a long tradition of empirical knowledge has developed, especially in Germany and Europe. Viscum album extract (VAE) is applied subcutaneously, normally two to three times per week whereas the complete treatment duration varies from some weeks up to five years and more. Different products are available such as ABNOBAViscum, Helixor, Iscador, or Lektinol.

Mistletoe contains biologically active molecules including lectins, flavonoides, viscotoxins, oligo- and polysaccharides, alkaloids, membrane lipids and other substances [4]. Although the exact pharmacological mode of action of mistletoe is not completely elucidated, there is a growing number of biological studies with a clear focus on lectins. Lectins (from the Latin *legere*, “to select”) are carbohydrate-binding proteins displayed on cell-surfaces to convey the interaction of cells with their environment [5]. Lectins mediate many immunological activities: For example, lectins show an immunomodulatory effect on neutrophils and macrophages by increasing the natural killer cytotoxicity and the number of activated lymphocytes [6-8]. They induce apoptosis in human lymphocytes [9] and boost the antioxidant system in mice [10]. In healthy subjects, the subcutaneous application of mistletoe has stimulated the production of granulocyte-macrophage colony-stimulating factor (GM-CSF), Interleukin 5 and Interferon gamma [11], indicating the immunopotentiating properties of mistletoe. The multiple ways how mistletoe affects the immune system have been recently reviewed elsewhere [12]. In consequence, the immunological pathways of conventional oncological treatments may be influenced by VAE, affecting cancer cells and decreasing adverse effects. This may result in a better quality of life (QoL).

A number of reviews has been published over the last two decades that address the effects of VAE on QoL in cancer patients [13-19]. However, these studies are either out of date, don’t make use of all published evidence, and/or don’t combine the data quantitatively into a pooled effect size. The aim of this study is therefore to review and analyze the current evidence regarding QoL of cancer patients which were treated with VAE and to calculate a meta-analysis.

## Methods

The study has been reported in accordance to PRISMA. The protocol was submitted to PROSPERO (registration number: CRD42019137704).

### Sources of evidence

We searched the databases Medline, Embase, PsychInfo, CENTRAL, CINAHL, Web of Science, and clinicaltrials.gov, we used google scholar, hand-searched the reference lists of reviews and identified studies and screened for grey literature via Google and opengrey.org. In case of missing data we contacted the authors.

### Search strategy

We developed a search strategy by iteratively combining synonyms and/or subterms of “quality of life” (e.g. well-being, QoL), “cancer” (e.g. neoplasm, sarcom, lymphom) and “mistletoe” (e.g. Helixor, Eurixor) to identify an adequate set of terms. We applied the following search strategy for Medline (Pubmed) and adopted it to the other databases accordingly:

1. quality of life OR HRQoL OR HRQL OR QOL OR patient satisfaction OR well-being OR wellbeing
2. mistel OR mistletoe OR Iscador OR Iscar OR Helixor OR Iscucin OR Abnobaviscum OR Eurixor OR Plenosol OR Lektinol OR Vysorel OR Isorel OR Cefalektin OR Viscum
3. Krebs OR cancer OR neoplasm/ OR tumor OR oncolog* OR onkologie OR carcin* OR malignant OR metastasis
4. Humans[MESH]
5. 1 AND 2 AND 3 AND 4

With the exception of #4 the general search fields were applied.

### Selection criteria

We included studies that measured QoL or self-regulation of cancer patients treated with mistletoe extracts assessed by performance status scales or patient-reported instruments. Studies were chosen if they were

- prospective controlled studies with
- two or more arms,
- both interventional and non-interventional.

The search was not limited to languages.

Studies were excluded if

- they did not meet the aforementioned inclusion criteria,
- if they tested multi-component complementary medicine interventions,
- if they failed to report sufficient information to be included into the meta-analysis or
- where this information cannot be gleaned from authors or extracted from graphs.

### Data management

The data was extracted from each study and entered into a spreadsheet by two authors independently. Then the extracted spreadsheets were compared and discrepancies were resolved by discussion until consensus was reached. We coded the following characteristics:

- number of participants in each treatment arm
- year, when study was conducted; in case this was not given, we estimated a 3 year lag from publication date for the meta-regression
- duration of study
- country where the study was conducted
- cancer type
- age
- gender of patients
- diagnosis according to ICD 10
- duration of study
- type of study (interventional vs. non-interventional, randomized vs. non-randomized, blinded vs. not blinded, single vs. multi-center)
- additional therapy (e.g. chemotherapy)
- number of drop-outs in each study arm
- active mistletoe extract preparation (e.g. Eurixor, Iscador, etc.)
- control treatment (e.g. placebo)
- effect size of primary outcomes plus standard deviation, or confidence intervals for effect measure provided using the reported global measure of QoL
- instrument used to measure primary outcomes
- statistics according to intent-to-treat analysis (yes/no)
- sponsoring of study (corporate, public, no-sponsoring).

If numerical data provided by the study publication was insufficient to calculate effect sizes, we contacted the authors. In cases where additional data were provided by the authors, these were then used instead of the published data. In older studies this was impossible. In those cases we used the given information (for instance means and confidence intervals, or means and p-values, or statistical information to generate the necessary data). In some cases we had to use medians as means and recover standard errors of the means from the given confidence intervals which also necessitated an adaptation of the confidence intervals into symmetrical ones. In each case we used the more conservative option which yielded larger standard errors and hence larger standard deviations. Thus, we generally opted for an error on the conservative side. When no quantitative information was given, but only graphs were presented, we printed high resolution graphs and derived the mean values and standard errors applying a ruler and used the given statistical information to arrive at the necessary quantitative scores. All these procedures were conducted independently and in duplicate [20].

### Risk of bias (quality) assessment

The Cochrane Risk of Bias tool 2 (Rob 2) was used to assess the risk of bias in randomized controlled trials [21]. All studies were assigned to the intention-to-treat-effect-analysis. Non-randomized or non-interventional studies were additionally analyzed with the Newcastle Ottawa Scale [22]. Two reviewers (HW, ML) independently assessed the risk of bias. In case of discrepancies they decided by consensus.

### Statistical analysis

The data were analyzed using Comprehensive Meta-Analysis V. 2 and Revman 5.3.5, the summary measure was the standardized mean difference. The meta-analysis was calculated independently by both authors using the two software tools Comprehensive Meta-Analysis and RevMan. The results were compared and underlying discrepancies resolved by discussion until both analyses yielded the same numerical results up to the second decimal. We report the overall analysis according to the results yielded by the RevMan analysis and conducted sensitivity analyses with Comprehensive Meta-Analysis.

The heterogeneity between studies was assessed by the Cochrane Q test and quantified by the index of heterogeneity (I^2^) [23]. A value of I^2^ of 25%, 50% and 75% indicates low, medium and high heterogeneity, respectively. If heterogeneity was higher than 25% we applied a random effects model for pooling the data, else a fixed effects model was used. As heterogeneity was high for the overall data-set, a random effects model was indicated. Fixed effect models were only used sparingly in exploratory subgroup analyses or sensitivity analyses, when heterogeneity was low.

We conducted subgroup analysis in order to identify possible sources of the heterogeneity. Stratified analyses were performed by: study types (e.g. blinded vs. not blinded, randomized versus non-randomized, types of control), additional treatments, country, risk-of-bias status, type of sponsoring, QoL instruments and related dimensions (in particular self-regulation), and mistletoe compound. Type of cancer was not included, as there were too many different cancer types. We conducted meta-regressions and regressed the three continuous predictors year of study, age of patients and length of treatment on effect size. We checked for publication bias using Egger’s regression intercept method and Duval and Tweedie’s trim and fill method [24].

## Results

598 studies were identified by electronic and hand searches, after removing duplicates. 67 full texts were retrieved of which 26 publications with 30 separate data sets met the inclusion criteria (see figure 1) [25-50]. We contacted 14 authors for additional information which was granted by five [25, 29, 39, 49, 51].

**Fig. 1:**
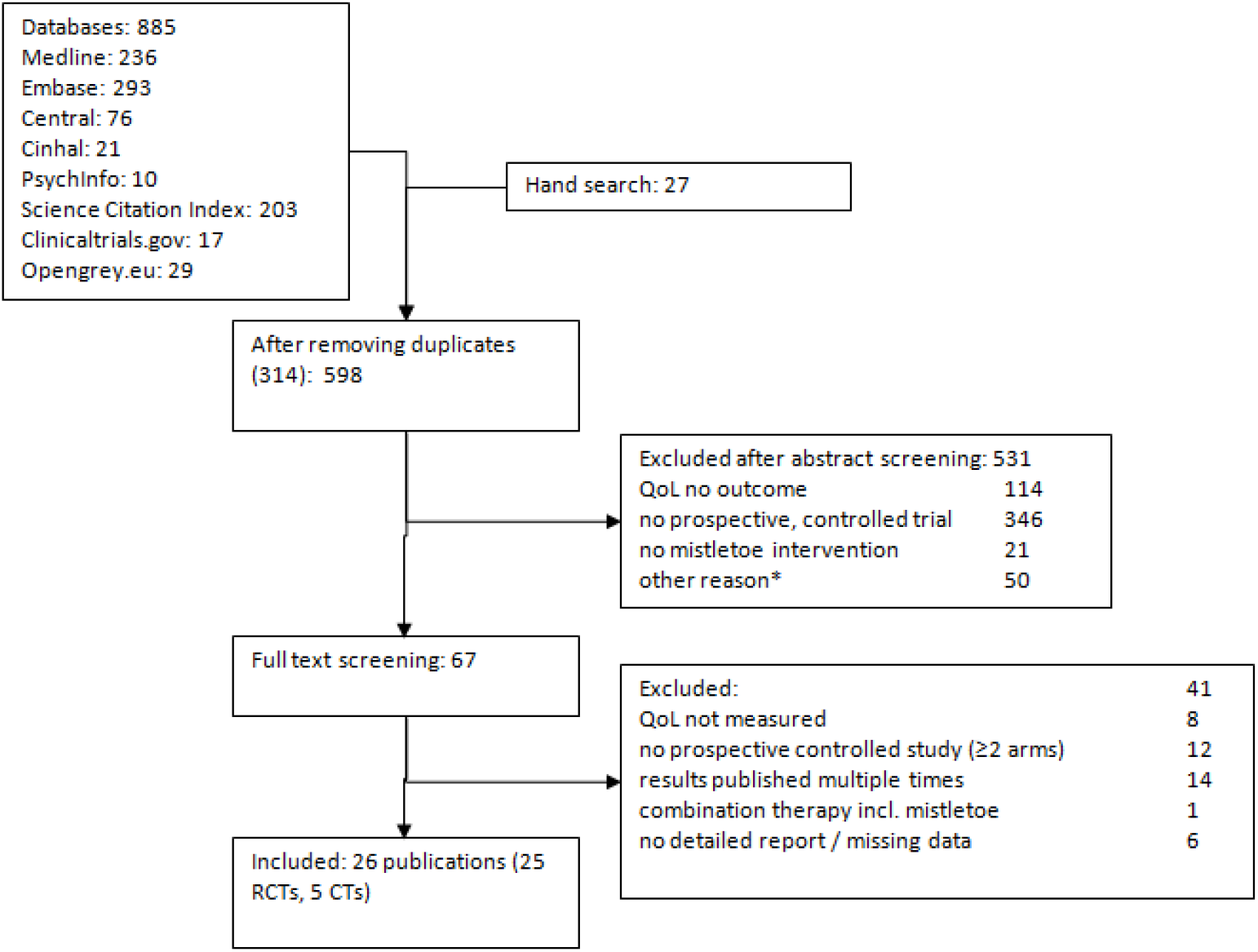
Flow of literature search process *e.g. not human, ongoing trials, finished trials without reports, results published multiple times

90% of the studies were conducted in Europe including Russia, 50% in Germany, and 10% in Asia. Three trials were blinded, four studies or datasets were not randomized. Different mistletoe preparations with varying conventional treatments were compared to conventional treatment (alone in 22 cases or plus an additional comparator in eight cases, respectively) for multiple types and stages of cancer. In nine studies QoL was measured with EORTC-QLQ-C30, six studies assessed self-regulation, and the others used one or multiple other instruments. The study characteristics are displayed in table 1.

**Table 1:**
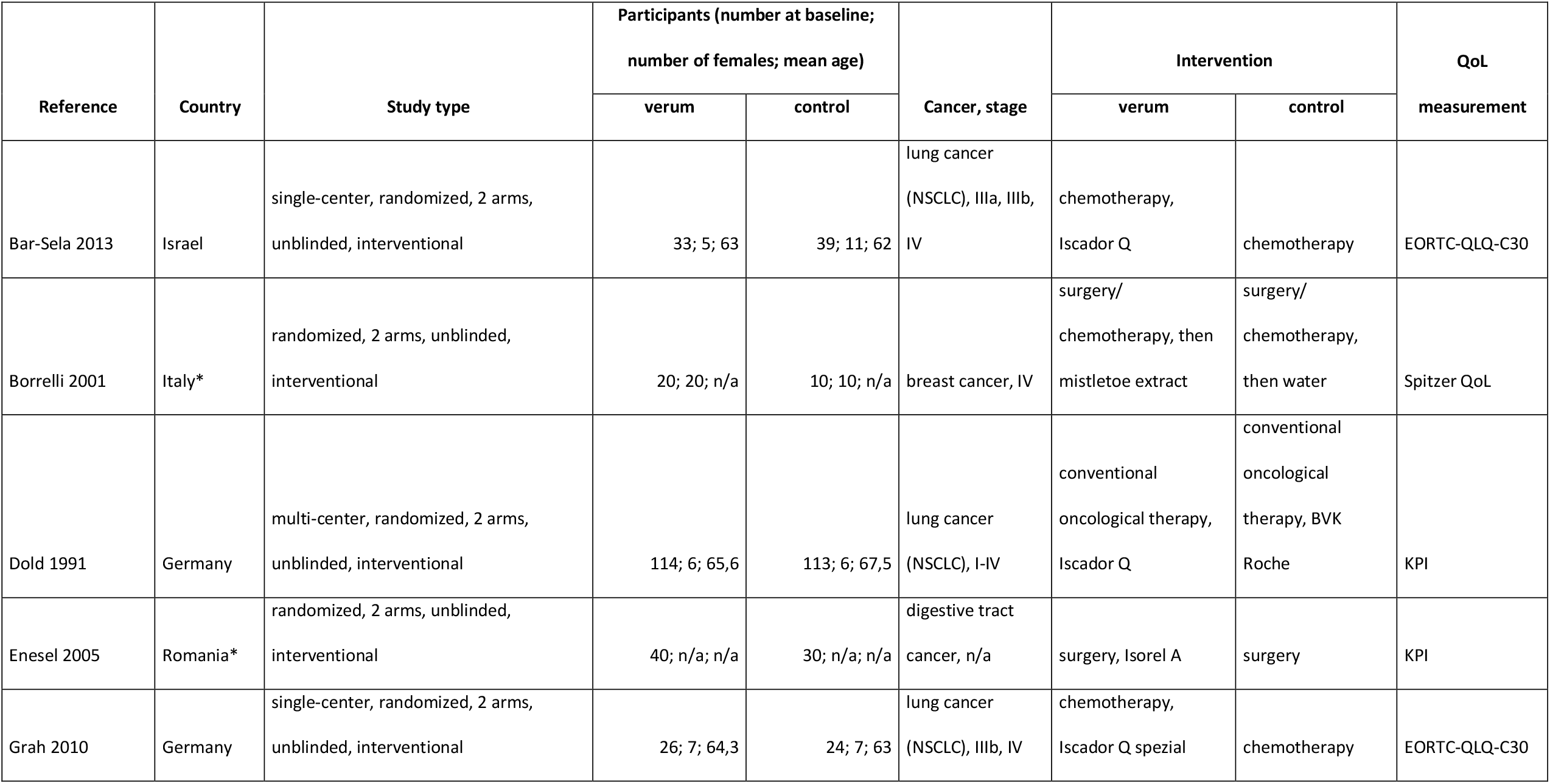
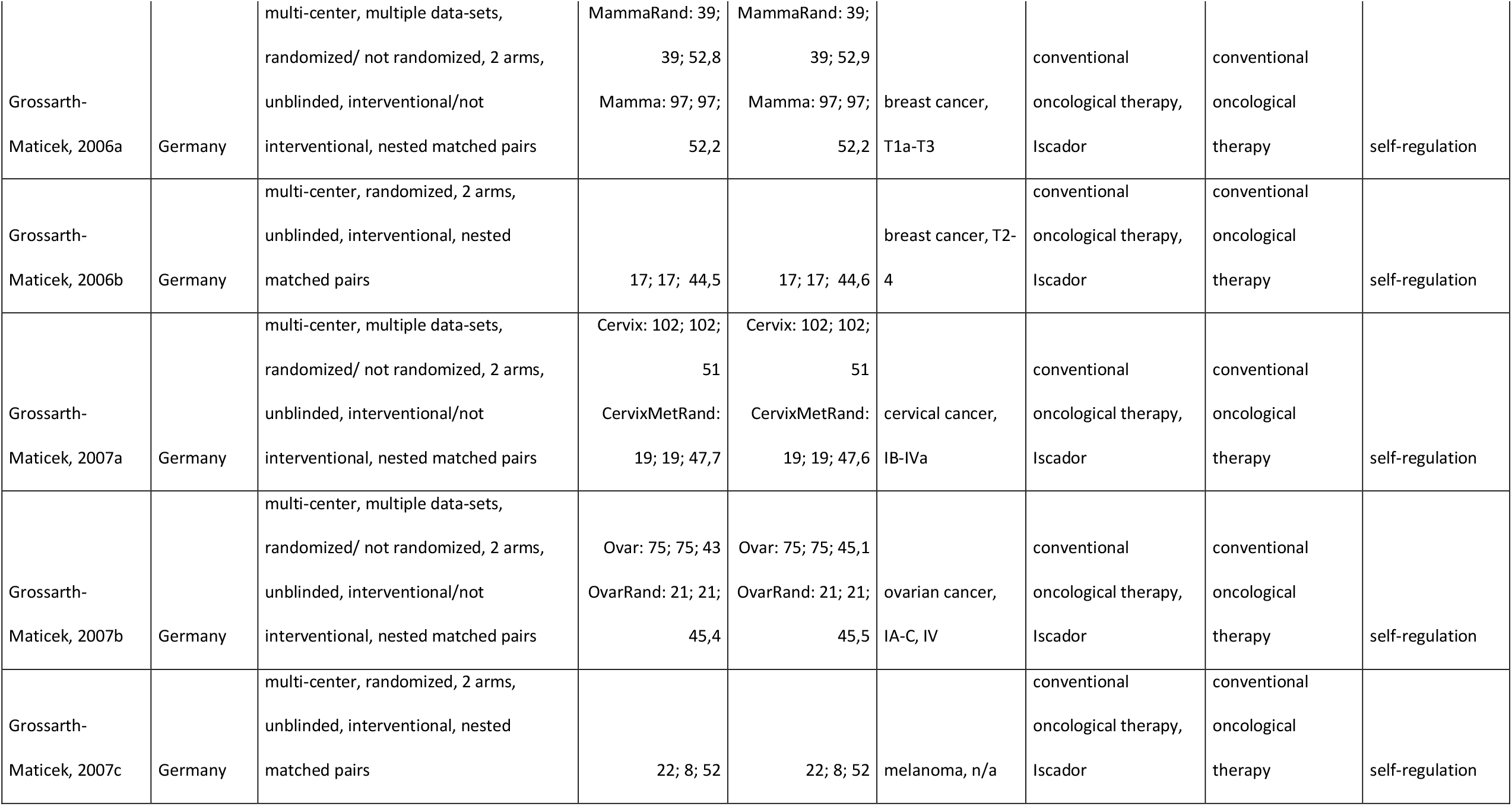
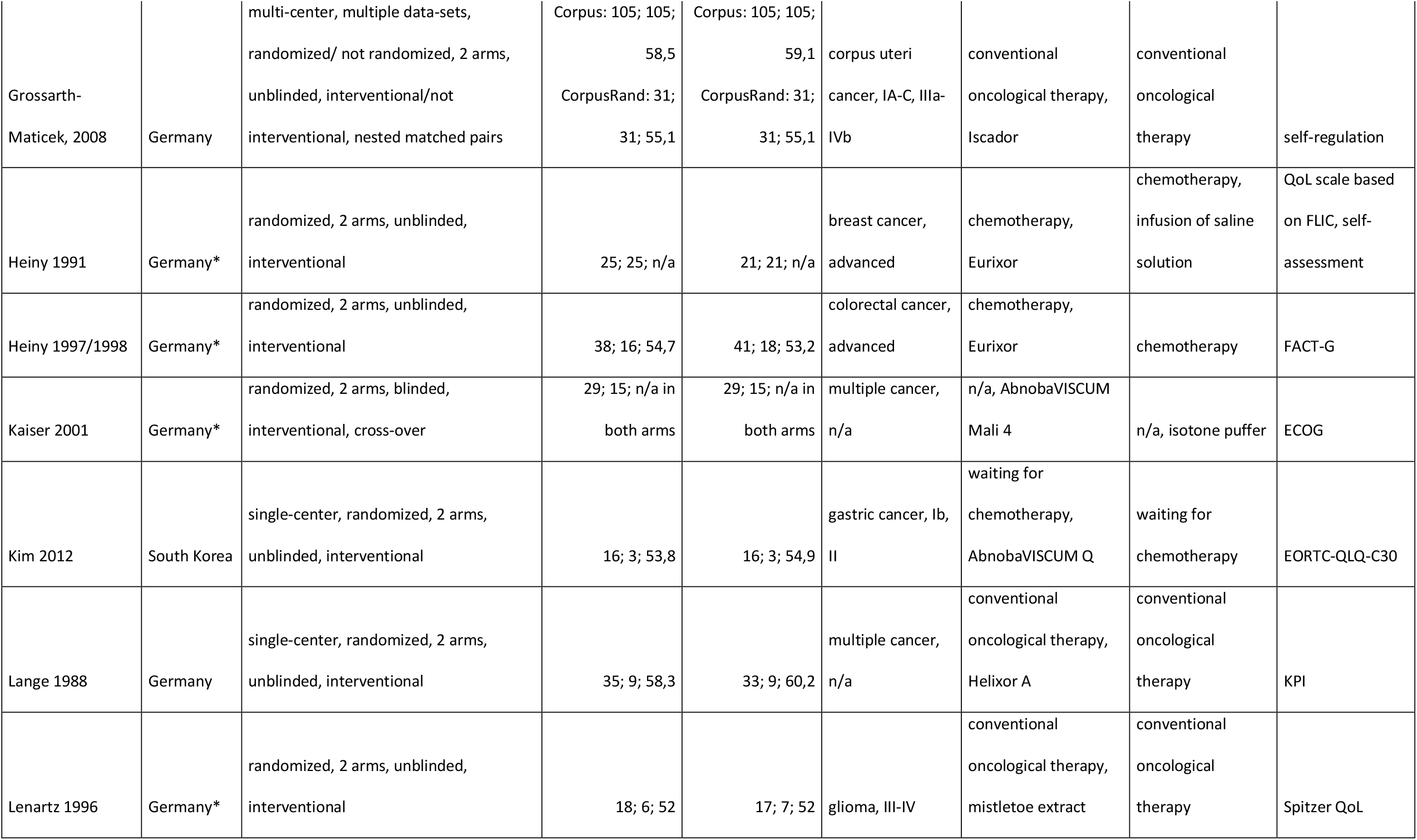
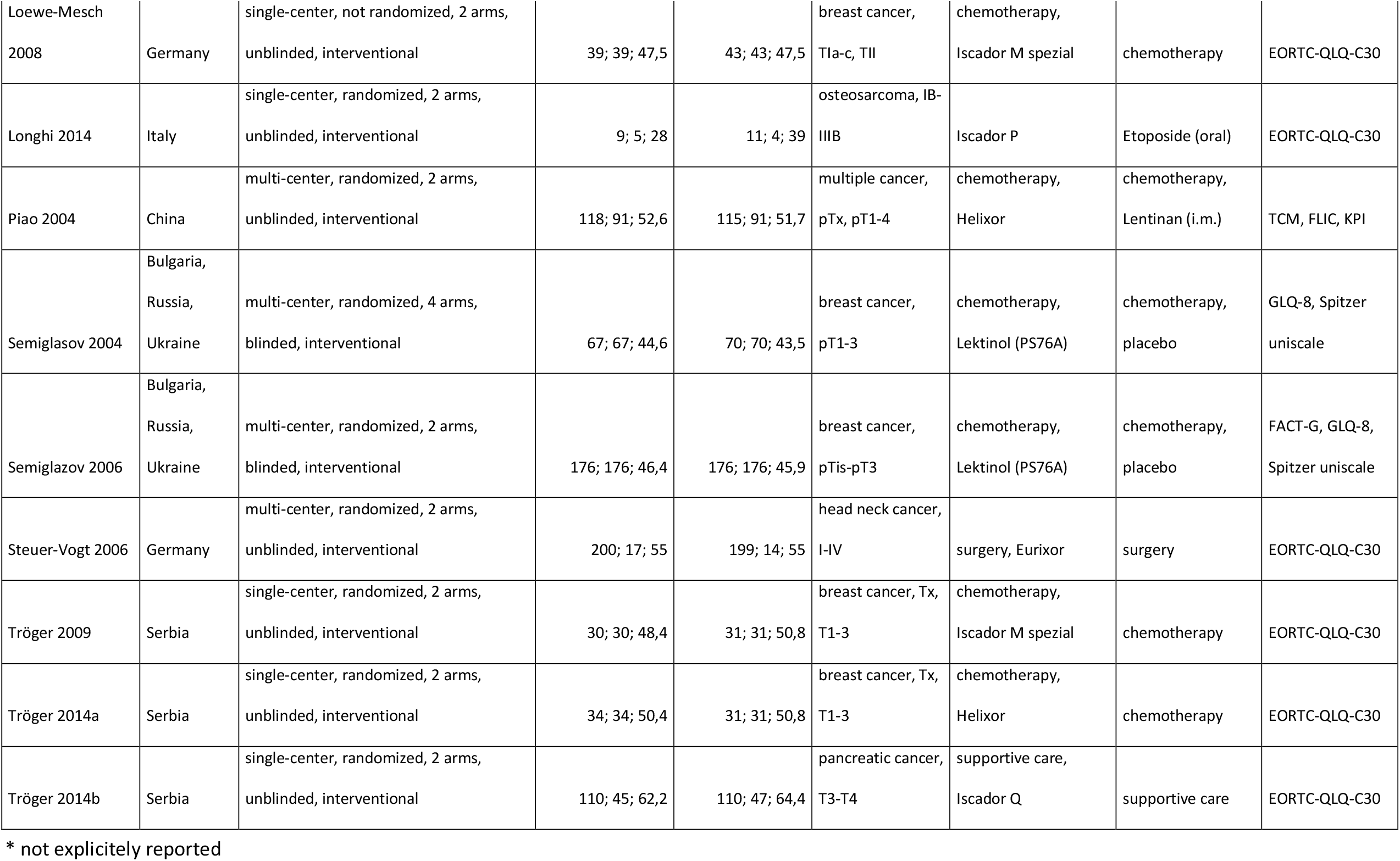
Characteristics of included studies

| Reference | Country | Study type | Participants (number at baseline;<br>number of females; mean age) |  | Cancer, stage | Intervention |  | QoL<br>measurement |
| --- | --- | --- | --- | --- | --- | --- | --- | --- |
|  |  |  | verum | control |  | verum | control |  |
| Bar-Sela 2013 | Israel | single-center, randomized, 2 arms,<br>unblinded, interventional | 33; 5; 63 | 39; 11; 62 | lung cancer<br>(NSCLC), IIIa, IIIb,<br>IV | chemotherapy,<br>Iscador Q | chemotherapy | EORTC-QLQ-C30 |
| Borrelli 2001 | Italy* | randomized, 2 arms, unblinded,<br>interventional | 20; 20; n/a | 10; 10; n/a | breast cancer, IV | surgery/<br>chemotherapy, then<br>mistletoe extract | surgery/<br>chemotherapy,<br>then water | Spitzer QoL |
| Dold 1991 | Germany | multi-center, randomized, 2 arms,<br>unblinded, interventional | 114; 6; 65,6 | 113; 6; 67,5 | lung cancer<br>(NSCLC), I-IV | conventional<br>oncological therapy,<br>Iscador Q | conventional<br>oncological<br>therapy, BVK<br>Roche | KPI |
| Enesel 2005 | Romania* | randomized, 2 arms, unblinded,<br>interventional | 40; n/a; n/a | 30; n/a; n/a | digestive tract<br>cancer, n/a | surgery, Isorel A | surgery | KPI |
| Grah 2010 | Germany | single-center, randomized, 2 arms,<br>unblinded, interventional | 26; 7; 64,3 | 24; 7; 63 | lung cancer<br>(NSCLC), IIIb, IV | chemotherapy,<br>Iscador Q spezial | chemotherapy | EORTC-QLQ-C30 |
| Grossarth-Maticek, 2006a | Germany | multi-center, multiple data-sets,<br>randomized/ not randomized, 2 arms,<br>unblinded, interventional/not<br>interventional, nested matched pairs | MammaRand: 39;<br>39; 52,8<br>Mamma: 97; 97;<br>52,2 | MammaRand: 39;<br>39; 52,9<br>Mamma: 97; 97;<br>52,2 | breast cancer,<br>T1a-T3 | conventional<br>oncological therapy,<br>Iscador | conventional<br>oncological<br>therapy | self-regulation |
| Grossarth-Maticek, 2006b | Germany | multi-center, randomized, 2 arms,<br>unblinded, interventional, nested<br>matched pairs | 17; 17; 44,5 | 17; 17; 44,6 | breast cancer, T2-4 | conventional<br>oncological therapy,<br>Iscador | conventional<br>oncological<br>therapy | self-regulation |
| Grossarth-Maticek, 2007a | Germany | multi-center, multiple data-sets,<br>randomized/ not randomized, 2 arms,<br>unblinded, interventional/not<br>interventional, nested matched pairs | Cervix: 102; 102;<br>51<br>CervixMetRand:<br>19; 19; 47,7 | Cervix: 102; 102;<br>51<br>CervixMetRand:<br>19; 19; 47,6 | cervical cancer,<br>IB-IVa | conventional<br>oncological therapy,<br>Iscador | conventional<br>oncological<br>therapy | self-regulation |
| Grossarth-Maticek, 2007b | Germany | multi-center, multiple data-sets,<br>randomized/ not randomized, 2 arms,<br>unblinded, interventional/not<br>interventional, nested matched pairs | Ovar: 75; 75; 43<br>OvarRand: 21; 21;<br>45,4 | Ovar: 75; 75; 45,1<br>OvarRand: 21; 21;<br>45,5 | ovarian cancer,<br>IA-C, IV | conventional<br>oncological therapy,<br>Iscador | conventional<br>oncological<br>therapy | self-regulation |
| Grossarth-Maticek, 2007c | Germany | multi-center, randomized, 2 arms,<br>unblinded, interventional, nested<br>matched pairs | 22; 8; 52 | 22; 8; 52 | melanoma, n/a | conventional<br>oncological therapy,<br>Iscador | conventional<br>oncological<br>therapy | self-regulation |
| Grossarth-Maticek, 2008 | Germany | multi-center, multiple data-sets, randomized/ not randomized, 2 arms, unblinded, interventional/not interventional, nested matched pairs | Corpus: 105; 105; 58,5<br>CorpusRand: 31; 31; 55,1 | Corpus: 105; 105; 59,1<br>CorpusRand: 31; 31; 55,1 | corpus uteri cancer, IA-C, IIIa-IVb | conventional oncological therapy, Iscador | conventional oncological therapy | self-regulation |
| Heiny 1991 | Germany* | randomized, 2 arms, unblinded, interventional | 25; 25; n/a | 21; 21; n/a | breast cancer, advanced | chemotherapy, Eurixor | chemotherapy, infusion of saline solution | QoL scale based on FLIC, self-assessment |
| Heiny 1997/1998 | Germany* | randomized, 2 arms, unblinded, interventional | 38; 16; 54,7 | 41; 18; 53,2 | colorectal cancer, advanced | chemotherapy, Eurixor | chemotherapy | FACT-G |
| Kaiser 2001 | Germany* | randomized, 2 arms, blinded, interventional, cross-over | 29; 15; n/a in both arms | 29; 15; n/a in both arms | multiple cancer, n/a | n/a, AbnobaVISCUM Mali 4 | n/a, isotone puffer | ECOG |
| Kim 2012 | South Korea | single-center, randomized, 2 arms, unblinded, interventional | 16; 3; 53,8 | 16; 3; 54,9 | gastric cancer, Ib, II | waiting for chemotherapy, AbnobaVISCUM Q | waiting for chemotherapy | EORTC-QLQ-C30 |
| Lange 1988 | Germany | single-center, randomized, 2 arms, unblinded, interventional | 35; 9; 58,3 | 33; 9; 60,2 | multiple cancer, n/a | conventional oncological therapy, Helixor A | conventional oncological therapy | KPI |
| Lenartz 1996 | Germany* | randomized, 2 arms, unblinded, interventional | 18; 6; 52 | 17; 7; 52 | glioma, III-IV | conventional oncological therapy, mistletoe extract | conventional oncological therapy | Spitzer QoL |
| Loewe-Mesch<br>2008 | Germany | single-center, not randomized, 2 arms,<br>unblinded, interventional | 39; 39; 47,5 | 43; 43; 47,5 | breast cancer,<br>T1a-c, TII | chemotherapy,<br>Iscador M spezial | chemotherapy | EORTC-QLQ-C30 |
| Longhi 2014 | Italy | single-center, randomized, 2 arms,<br>unblinded, interventional | 9; 5; 28 | 11; 4; 39 | osteosarcoma, IB-IIIB | Iscador P | Etoposide (oral) | EORTC-QLQ-C30 |
| Piao 2004 | China | multi-center, randomized, 2 arms,<br>unblinded, interventional | 118; 91; 52,6 | 115; 91; 51,7 | multiple cancer,<br>pTx, pT1-4 | chemotherapy,<br>Helixor | chemotherapy,<br>Lentinan (i.m.) | TCM, FLIC, KPI |
| Semiglasov 2004 | Bulgaria,<br>Russia,<br>Ukraine | multi-center, randomized, 4 arms,<br>blinded, interventional | 67; 67; 44,6 | 70; 70; 43,5 | breast cancer,<br>pT1-3 | chemotherapy,<br>Lektinol (PS76A) | chemotherapy,<br>placebo | GLQ-8, Spitzer<br>uniscale |
| Semiglazov 2006 | Bulgaria,<br>Russia,<br>Ukraine | multi-center, randomized, 2 arms,<br>blinded, interventional | 176; 176; 46,4 | 176; 176; 45,9 | breast cancer,<br>pTis-pT3 | chemotherapy,<br>Lektinol (PS76A) | chemotherapy,<br>placebo | FACT-G, GLQ-8,<br>Spitzer uniscale |
| Steuer-Vogt 2006 | Germany | multi-center, randomized, 2 arms,<br>unblinded, interventional | 200; 17; 55 | 199; 14; 55 | head neck cancer,<br>I-IV | surgery, Eurixor | surgery | EORTC-QLQ-C30 |
| Tröger 2009 | Serbia | single-center, randomized, 2 arms,<br>unblinded, interventional | 30; 30; 48,4 | 31; 31; 50,8 | breast cancer, Tx,<br>T1-3 | chemotherapy,<br>Iscador M spezial | chemotherapy | EORTC-QLQ-C30 |
| Tröger 2014a | Serbia | single-center, randomized, 2 arms,<br>unblinded, interventional | 34; 34; 50,4 | 31; 31; 50,8 | breast cancer, Tx,<br>T1-3 | chemotherapy,<br>Helixor | chemotherapy | EORTC-QLQ-C30 |
| Tröger 2014b | Serbia | single-center, randomized, 2 arms,<br>unblinded, interventional | 110; 45; 62,2 | 110; 47; 64,4 | pancreatic cancer,<br>T3-T4 | supportive care,<br>Iscador Q | supportive care | EORTC-QLQ-C30 |

\* not explicitly reported

The results of the overall meta-analysis are presented in Figure 2.

**Fig. 2:**
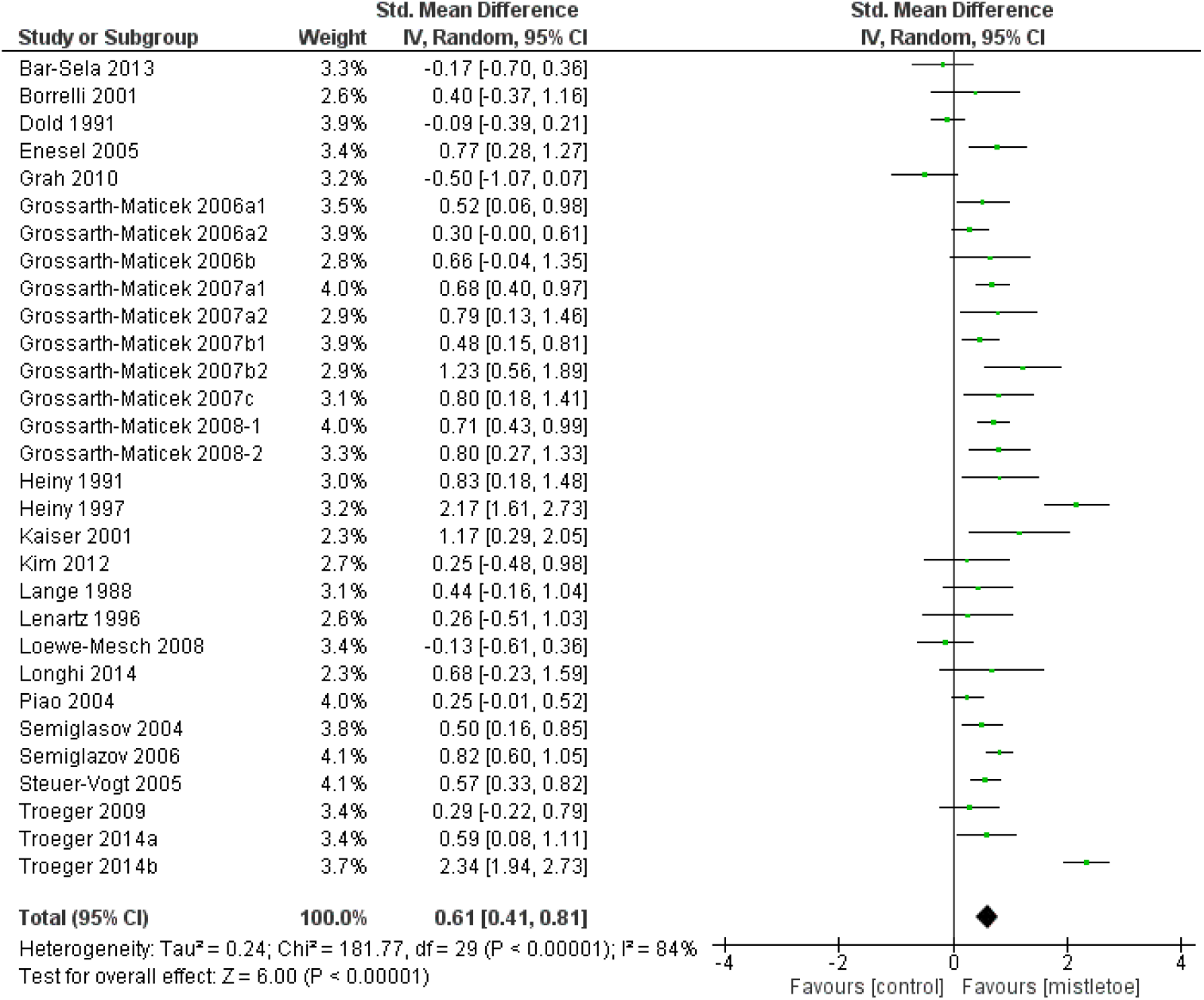
Overall Meta-analysis of all included data sets

As can be seen the studies are highly heterogeneous (I^2^ = 84%), and hence the random effects model is applied to estimate the combined standardized mean difference as d = 0.61 (95% CI 0.41-0.81, p<0.00001, z=6.05).

The meta-analyses of the sub-dimensions of QoL are shown in table 2. The SMD of seven out of 14 QoL dimensions are significant (p≤0.05). The pooled SMD of role and social functioning are 0.63 (95% CI 0.05-1.22) and 0.62 (95% CI 0.22-1.03), respectively. For pain, it is SMD=-0.86 (95% CI -1.54-(-0.18)) and for nausea, it is SMD=-0.55 (95% CI -1-(-0.1)).

**Table 2:**
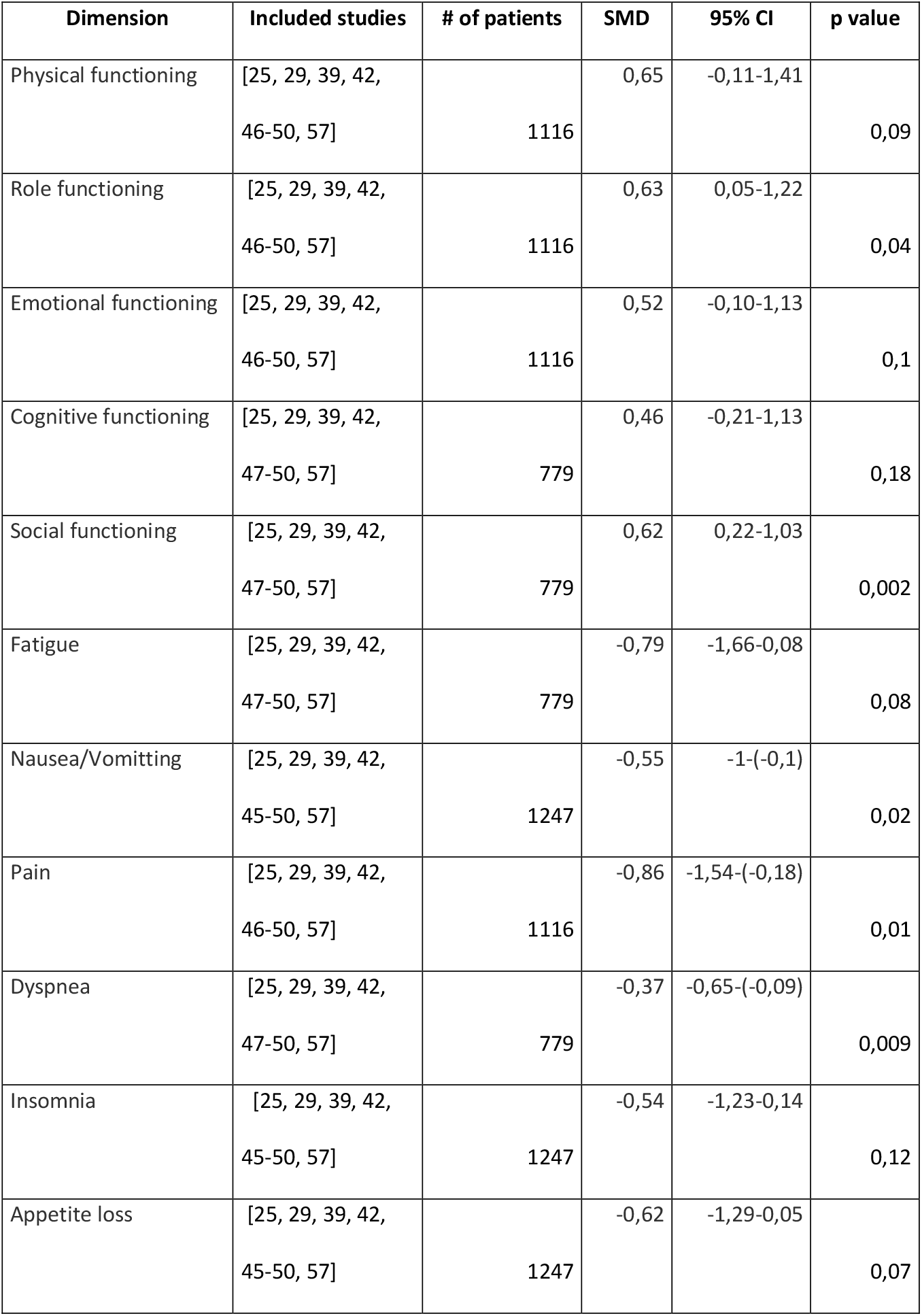
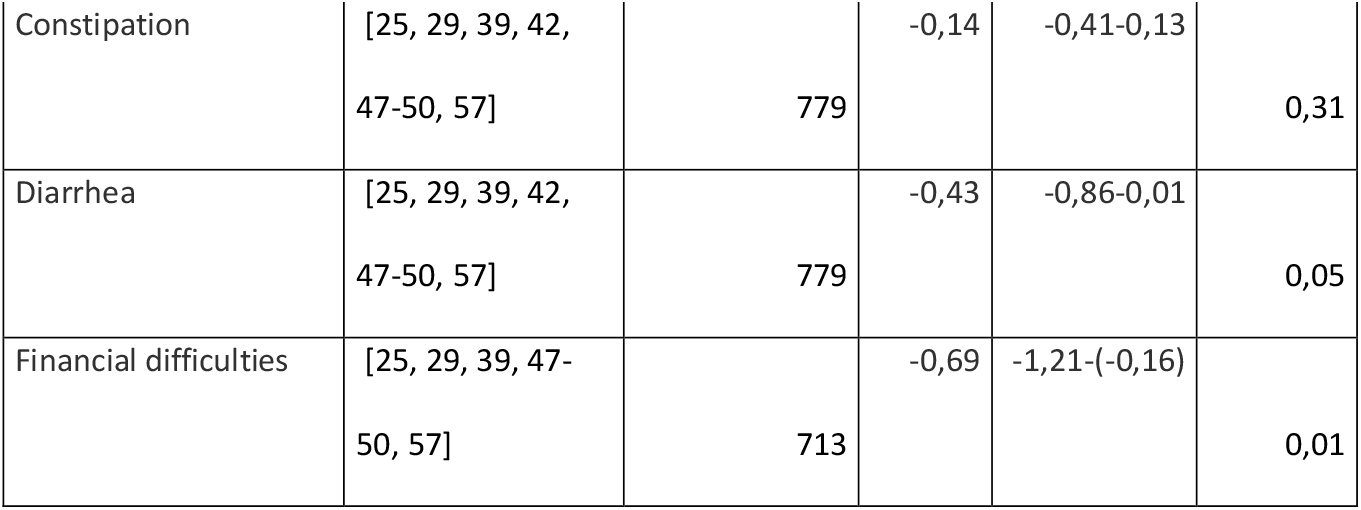
Effect sizes of sub-dimensions of QoL that could be pooled by meta-analyses (positive [negative] values in functioning [symptom] dimensions indicate improvement for VAE vs. control)

| Dimension | Included studies | # of patients | SMD | 95% CI | p value |
| --- | --- | --- | --- | --- | --- |
| Physical functioning | [25, 29, 39, 42,<br>46-50, 57] | 1116 | 0,65 | -0,11-1,41 | 0,09 |
| Role functioning | [25, 29, 39, 42,<br>46-50, 57] | 1116 | 0,63 | 0,05-1,22 | 0,04 |
| Emotional functioning | [25, 29, 39, 42,<br>46-50, 57] | 1116 | 0,52 | -0,10-1,13 | 0,1 |
| Cognitive functioning | [25, 29, 39, 42,<br>47-50, 57] | 779 | 0,46 | -0,21-1,13 | 0,18 |
| Social functioning | [25, 29, 39, 42,<br>47-50, 57] | 779 | 0,62 | 0,22-1,03 | 0,002 |
| Fatigue | [25, 29, 39, 42,<br>47-50, 57] | 779 | -0,79 | -1,66-0,08 | 0,08 |
| Nausea/Vomitting | [25, 29, 39, 42,<br>45-50, 57] | 1247 | -0,55 | -1-(-0,1) | 0,02 |
| Pain | [25, 29, 39, 42,<br>46-50, 57] | 1116 | -0,86 | -1,54-(-0,18) | 0,01 |
| Dyspnea | [25, 29, 39, 42,<br>47-50, 57] | 779 | -0,37 | -0,65-(-0,09) | 0,009 |
| Insomnia | [25, 29, 39, 42,<br>45-50, 57] | 1247 | -0,54 | -1,23-0,14 | 0,12 |
| Appetite loss | [25, 29, 39, 42,<br>45-50, 57] | 1247 | -0,62 | -1,29-0,05 | 0,07 |
| Constipation | [25, 29, 39, 42,<br>47-50, 57] | 779 | -0,14 | -0,41-0,13 | 0,31 |
| Diarrhea | [25, 29, 39, 42,<br>47-50, 57] | 779 | -0,43 | -0,86-0,01 | 0,05 |
| Financial difficulties | [25, 29, 39, 47-50, 57] | 713 | -0,69 | -1,21-(-0,16) | 0,01 |

The risk of bias assessment is displayed in the figures 3 and 4. 65% had an overall high risk of bias which resulted for most studies from the 85% high risk of bias in the measurement of outcome. This can be attributed to the missing blinding process, the QoL assessment as patient-reported outcome, and the uncertain appropriateness of some measurement instruments which may only incompletely capture the concept of QoL.

**Fig. 3:**
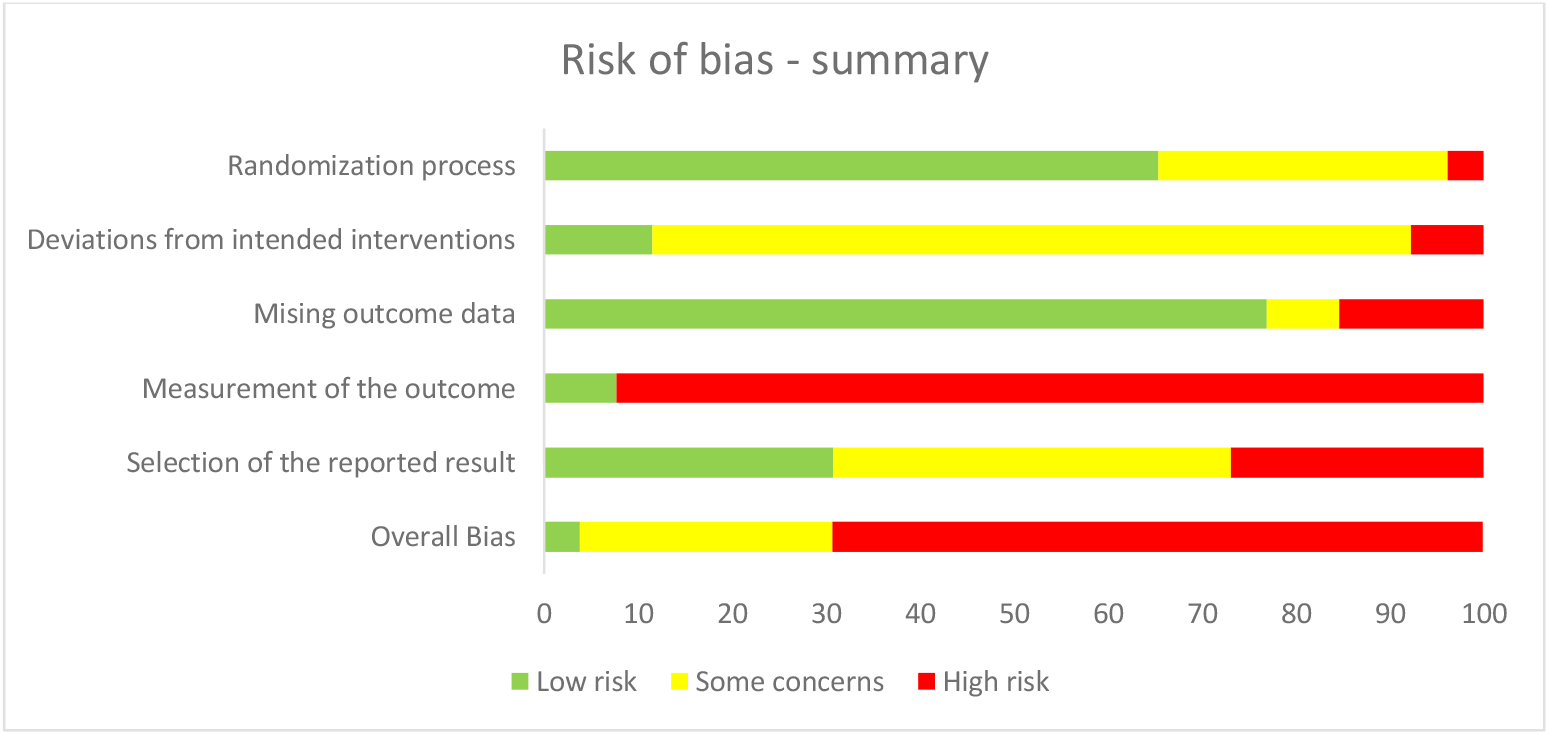
Summary of risk of bias assessment as percentage (intention-to-treat)

**Fig. 4:**
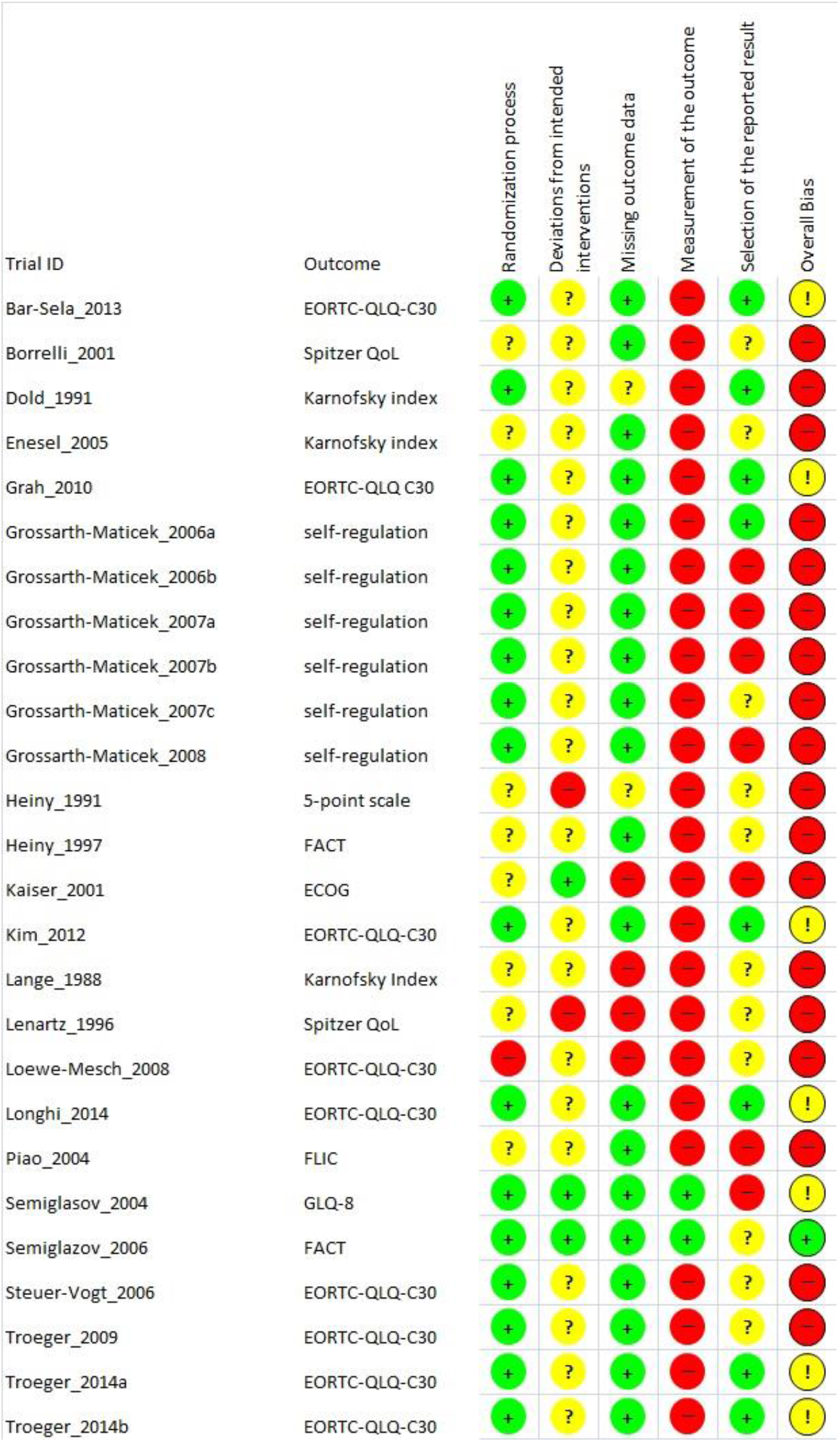
Risk of bias assessment by domain and overall bias

The five non-randomized trials [30, 33-35, 42] were additionally assessed with the Newcastle-Ottawa-scale. All studies had an overall score of 7 out of a maximum of 9. The sums in the selection, the comparability and the outcome/exposure domain were 3, 2 and 2, respectively, for all studies.

The sensitivity analyses are presented in Table 3.

**Table 3:** Sensitivity analyses according to various moderators

| **Moderator** | **N studies** | **Effect**<br>**Sizes**<br>**SMD\*** | **95% CIs** | **Heterogeneity**<br>**I<sup>2</sup>** | **z-score** | **p value <=§** |
| --- | --- | --- | --- | --- | --- | --- |
| *Risk of Bias status* |  |  |  |  |  |  |
| High | 22 | 0.66 | 0.41-0.90 | 75.5 | 5.2 | 0.001 |
| Low | 1 | 0.84 | 0.62-1.1 | 0 | 7.4 | 0.001 |
| Some | 7 | 0.56 | 0.12-1.0 | 93.7 | 2.5 | 0.014 |
| *Blinding* |  |  |  |  |  |  |
| Yes | 3 | 0.96 | 0.30-1.62 | 77.4 | 2.9 | 0.041 |
| No | 27 | 0.61 | 0.39-0.82 | 85.3 | 5.4 | 0.001 |
| *Randomized* |  |  |  |  |  |  |
| Yes | 25 | 0.70 | 0.47-0.93 | 86.3 | 6.0 | 0.001 |
| No | 5 | 0.38 | -0.09-0.85 | 63.4 | 1.58 | 0.11 |
| *Additional Treatment* |  |  |  |  |  |  |
| Chemotherapy | 7 | 0.41 | 0.05-0.76 | 89.3 | 2.2 | 0.025 |
| No add. Treatment | 4 | 0.77 | 0.20-1.34 | 64.6 | 2.6 | 0.008 |
| Individual best care | 1 | 2.33 | 1.93-3.38 | 0 | 5.2 | 0.001 |
| Conventional | 16 | 0.58 | 0.36-0.81 | 66.2 | 5.1 | 0.001 |
| Surgery | 2 | 0.62 | 0.40-0.83 | 0 | 5.5 | 0.001 |
| *Controls* |  |  |  |  |  |  |
| Active | 8 | 0.60 | 0.20-1.01 | 81.4 | 2.9 | 0.004 |
| No active | 22 | 0.65 | 0.41-0.90 | 85.9 |  | 0.001 |
| *Cancer type* |  |  |  |  |  |  |
| Lung cancer | 3 | -0.18 | -0.41-0.06 | 0 | 1.45 | 0.15 |
| Breast cancer | 10 | 0.48 | 0.29-0.68 | 50 | 4.83 | 0.00001 |
| *Product* |  |  |  |  |  |  |
| Abnova V. | 2 | 1.06 | 0.07-2.04 | 86.5 | 2.1 | 0.036 |
| Eurixor | 4 | 0.94 | 0.32-1.60 | 87.7 | 3.0 | 0.003 |
| Helixor | 3 | 0.35 | 0.13-0.57 | 0 | 3.1 | 0.002 |
| Iscador | 17 | 0.58 | 0.28-0.87 | 88.6 | 3.8 | 0.001 |
| Lektinol | 2 | 0.67 | -0.13-1.48 | 63.4 | 1.6 | 0.1 |
| Other | 2 | 0.67 | 0.26-1.09 | 0 | 3.2 | 0.001 |
| *Country* |  |  |  |  |  |  |
| Germany | 19 | 0.64 | 0.38-0.90 | 80.5 | 4.8 | 0.001 |
| Other | 11 | 0.64 | 0.30-0.98 | 89.4 | 3.7 | 0.001 |
| *Sponsoring* |  |  |  |  |  |  |
| Corporate | 10 | 0.49 | 0.11-0.87 | 92.0 | 2.5 | 0.011 |
| Public | 3 | 0.64 | -0.05-1.33 | 91.1 | 1.8 | 0.07 |
| Mixed | 10 | 0.73 | 0.36-1.1 | 35.2 | 3.8 | 0.001 |
| No Information | 7 | 0.73 | 0.27-1.19 | 82.8 | 3.1 | 0.002 |
| *Type of Measure* |  |  |  |  |  |  |
| Index<sup>1</sup> | 6 | 0.33 | -0.12-0.78 | 50.1 | 1.4 | 0.1 |
| Scale<sup>2</sup> | 14 | 0.71 | 0.41-1.0 | 90.9 | 4.6 | 0.001 |
| Self Regulation<sup>3</sup> | 10 | 0.73 | 0.39-1.07 | 35.2 | 4.1 | 0.001 |

\*if heterogeneity > 25 random effects SMDs are given, else fixed effect
\$ two-tailed
1 Karnofsky Index, ECOG, Spitzer QoL
2 EORTC QoL Q30, FACT, GLQ-8
3 Grossarth-Maticek's self-regulation scale

The sensitivity analyses confirmed the robustness of the results. Neither the methodological nor other moderator variables showed strong deviations. With the exception of the non-randomized studies (non-randomized: d = 0.38, p = 0.1), the lung cancer studies (d=-0.18, p = 0.15), the studies conducted with Lektinol (d = 0.67, p = 0.1), and the studies using an index measure (e.g. Karnofsky index) as outcome (d = 0.33, p = 0.1) all other moderator analyses showed no appreciable differences between subgroups and yielded highly significant effect sizes. In tendency, methodologically more rigorous studies yielded higher or equally high effect sizes than less rigorous ones. Most notably, randomized studies yielded a higher effect size (d = 0.70, p = 0.001) than non-randomized ones (d = 0.38, p = 0.1). Studies using active controls (d = 0.6, p = 0.004) did not differ from studies using other controls (d = 0.65, p < 0.001). Various types of additional treatment did not show differential effect sizes, except individualized best care, which, however, is an estimate based on only one study and hence not reliable. Although the effect sizes of the various products vary, their confidence intervals overlap, and hence suggest the conclusion that they are roughly equally effective. There is no difference in effect sizes depending on countries, type of sponsoring, or type of measures. Studies that relied on corporate sponsoring, and studies using only a single index measure yielded a somewhat smaller effect size, although confidence intervals overlap and thus signal non-significant differences.

The three meta-regressions are presented in Table 4 and in Figures 5 and 6.

**Table 4:** Meta-regression results

| Moderator | Point estimate | 95% CIs | z-score | p value <= |
| --- | --- | --- | --- | --- |
| *Study Year* |  |  |  | Model: p = 0.0001 |
| Slope | 0.026 | 0.012-0.04 | 3.8 | 0.001 |
| *Age* |  |  |  | Model: p = 0.3 |
| Slope | -0.006 | -0.02-0.006 | -1.05 | 0.2 |
| *Duration of Treatment* |  |  |  | Model: p = 0.06 |
| Slope | 0.04 | -0.0003-0.008 | 1.8 | 0.07 |

**Fig. 5:**
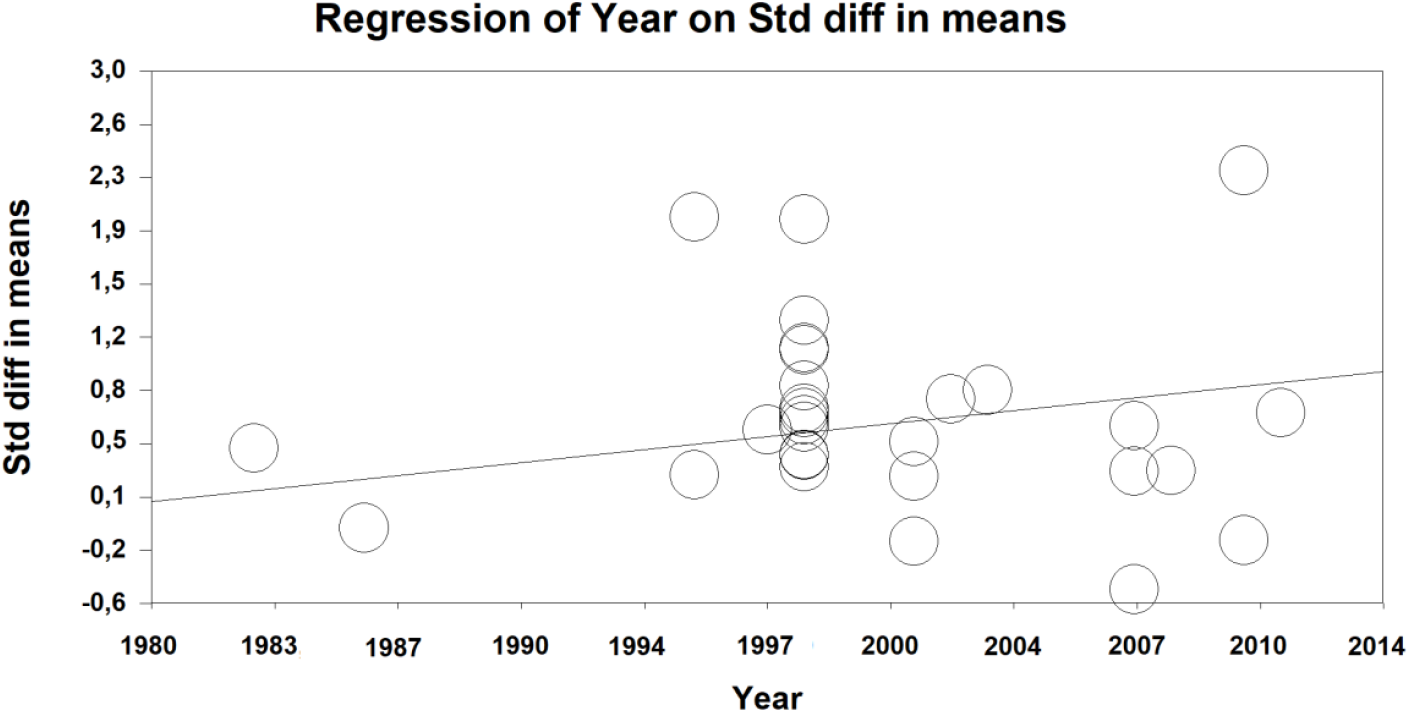
Scatterplot of the meta-regression of study year on effect-size

**Fig. 6:**
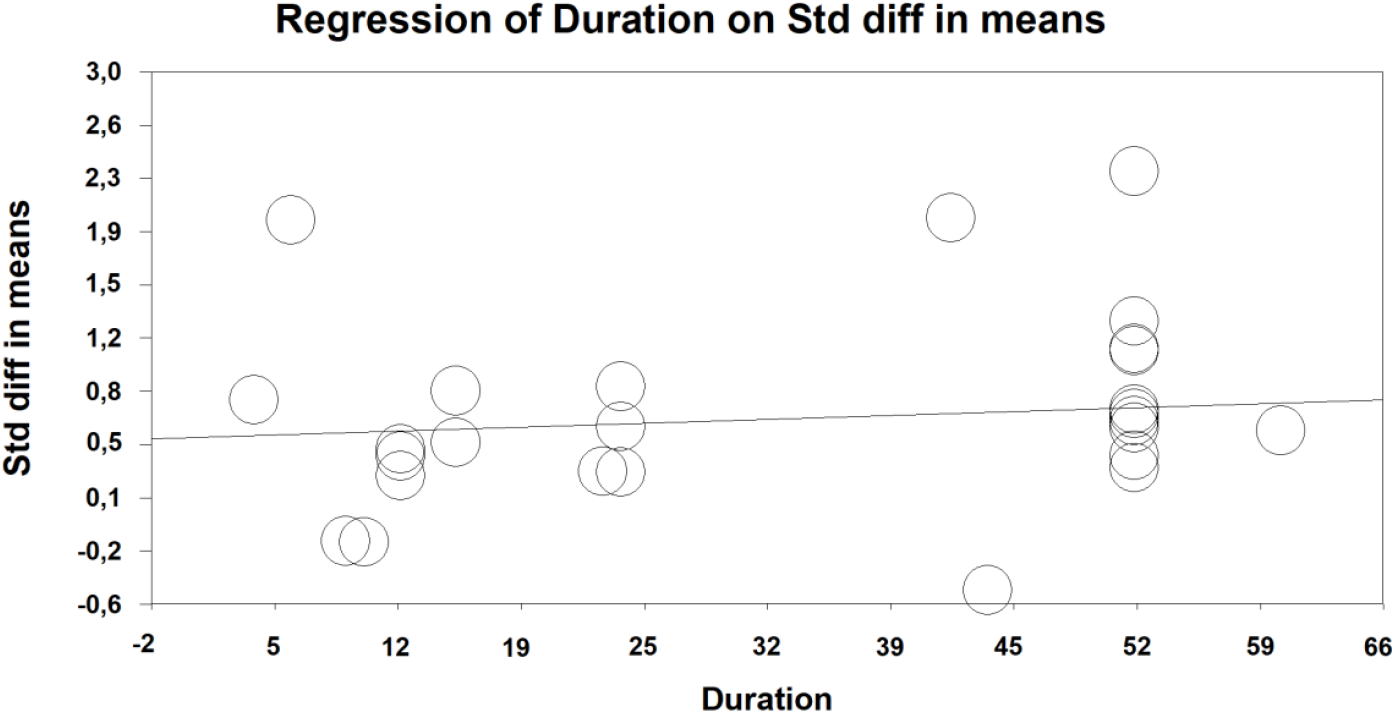
Scatterplot of the meta-regression of treatment duration on effect size

The study year is positively correlated with effect size. For each year the study was more recent the estimated QoL-effect size is larger by d = 0.03. Although there is a tendency for a larger effect size in younger patients this effect is not significant. The slope of the regression line for the duration of treatment is borderline significant, indicating that longer treatment produces effects that are 0.04 standard deviations larger per additional treatment week. Note though that only treatments between 5 and 52 weeks have entered the analysis and the variance is not large.

Publication bias was estimated using two methods. Egger’s regression intercept model regresses effect size on precision of study with the assumption that smaller studies that are less precise will more often go unpublished. A regression line with a lot of smaller and more imprecise studies missing should thus miss the origin by a large margin. In our analysis the intercept of the regression is 0.82 with a non-significant deviation from the origin (t = 0.65, p-value two tailed = 0.5). Duval and Tweedie’s trim and fill method is an extension of the graphical funnel plot analysis and analyzes how many studies would have to be trimmed to generate a perfectly symmetrical funnel plot. In our analysis this method estimates no studies to be trimmed on the left side, i.e on the negative or low side of the effect size estimate and an estimate of 7 trimmed studies on the right side, i.e. on the positive side of the effect size funnel with an adjustment that leads to a higher effect size, if the studies are trimmed. These two analyses of publication bias show that publication bias is not a likely explanation of this result and any funnel asymmetries are not due to unpublished studies but due to positive outliers.

## Discussion

This meta-analysis shows a significant and robust medium-sized effect of d = 0.61 of viscum album extract (VAE) treatment on QoL in cancer patients.

The results should be regarded in the light of the following facts:

The included studies vary with regard to the cancer site, the control intervention, the additional oncological treatment, and the VAE. While sensitivity analyses confirmed the robustness and reliability of the findings, they could not account for the heterogeneity of the effect sizes. Neither methodological moderators (blinded vs. unblinded studies, randomized versus non-randomized, studies with high versus low risk of bias, active versus non-active control) nor structural moderators (type of outcome measure, funding, VAE product used, additional treatment) could clarify the heterogeneity. We suspect that this is due to multiple interactions between cancer types and stages, treatments and structural variables that cannot be explored with a limited set of 30 studies. Nevertheless, our sensitivity analyses document the overall robustness of the effect, as none of the levels of moderators exhibits significant deviations from other levels or from the overall effect size. This gives our effect size estimate of d = 0.61 reliability.

Although the overall risk of bias is high in many studies, one should bear in mind two aspects. First, we applied the intention-to-treat-algorithm of Rob2 as the more conservative approach and not the per-protocol evaluation which may have resulted in a better overall bias. Second, due to the local skin reaction of VAE application the blinding of participants and carers is practically impossible and could only have been implemented reliably with an active placebo, which is ethically questionable. In Rob2 this leads to a high risk of bias in the measurement of the outcome. On the one hand, the lack of blinding might have biased the results since most QoL are self-reported and there may be strong beliefs among users of anthroposophic medicine which might additionally be fortified by the severity of the disease and the hope that an additional treatment has a positive impact [52]. It was shown that these attitudes are correlated with a better QoL [53]. On the other hand, there is no evidence from the included studies that the attitudes differed between treatment arms and if patients in the control group searched and used for surrogate medications for the VAE, this bias would favor the comparator. Furthermore, our sensitivity analysis gives no indication that studies with blinding and without blinding estimate different effect sizes and there is also no difference in effect sizes between studies from Germany – where mistletoe is well known – and other countries where mistletoe is less known and used. The results of the Newcastle-Ottawa-Scale, finally, indicate a good methodological quality for the non-randomized trials that were included in the review.

Another limitation is that self-regulation, the Karnofsky performance index, or the ECOG scale cover important aspects of QoL, but are different in content from other measures such as the global QoL of the EORTC-QLQ C30 scale. This source of heterogeneity was also addressed by our sensitivity analyses. This showed that, indeed, as one would expect, single item indices estimate lower effect sizes, although the difference is not significant. In the same vein, the inclusion of non-randomized and non-interventional trials might have biased the results due to their lower internal validity, but their exclusion during sensitivity analyses again did not alter the significance of the pooled outcome. In addition, four of the five non-RCTs had a matched-pair design which increases the comparability between treatment arms compared to other types of group allocation.

The meta-regression shows that more recent studies have higher effect sizes compared to older studies. This is counterintuitive at first sight, as normally more recent studies are implemented with more methodological rigor due to the GCP guidelines and a higher methodological skill of trialists. This, one would think, should, if at all, lead to smaller effect sizes in more recent trials. The fact that this is not the case shows, together with our sensitivity analysis that methodological bias is an unlikely explanation for the effect size found. However, another point is worth bearing in mind: earlier studies were very often implemented with severely ill patients with tumor status IV or in palliative care. Only in more recent studies was VAE also used as add on treatment in first line patients with a relatively good chance of surviving. Thus the higher effect size for more recent studies might also reflect the less severe status of these patients.

Our review has a number of strengths. First, we conducted a comprehensive search for published and grey literature with no time or language limitation to minimize publication bias. Our analysis of publication bias supports the conclusion that the effect size estimate is not due to publication bias. Some authors who we contacted, however, failed to provide additional information and the respective studies were consequently excluded. Second, we calculated a pooled SMD for a global measure of QoL and for its subdomains such as pain or fatigue. Third, we analyzed the data both with Revman 5.3 and CMA software which implements the Hunter-Schmidt-corrections for small sample bias. We did both analyses in parallel and independently, thus preventing coding or typing errors from biasing our results.

The weaknesses of this review are obvious. Any meta-analysis can only be as good as the original studies entered. Some of these studies are large and methodologically strong. But some are also badly reported, small and with a mixed patient load. In some cases we had to recalibrate confidence interval estimates, because the data given were not detailed enough. Although it would have been desirable, the variance between cancer types and stages was too large to allow for detailed assessments and separate analyses, which might have reduced the heterogeneity. Although we can testify to the robustness of the overall effect size estimate, we have not succeeded in clarifying the heterogeneity of the studies. This requires multi-center studies in large cohorts of patients with large budgets. Thus, one consequence of this meta-analysis would be to call for more serious efforts from public funders to study the effects of VAEs in large and homogeneous patient cohorts to confirm or disconfirm the results of this analysis.

### Clinical relevance

Our results indicate a statistically significant and clinically valuable improvement of the subjective well-being of patients with different types of cancer after the treatment with VAE. The analyses for the subdomains revealed a significant pooled SMD for important symptoms and functioning indices, whereas other show a positive, yet not significant effect of VAE compared to control. Whether these vital elements of QoL such as emotional functioning or fatigue are influenced remain statistically uncertain. Overall, a robust estimate of an improvement of d = 0.61 in quality of life represents a medium-sized [54] and clinically relevant [55, 56] effect that makes VAE treatment a viable add-on option to any anticancer treatment.

### Conclusion

Our analysis provides evidence that global QoL in cancer patients is positively influenced by VAE. Because the risk of bias and the heterogeneity is high, future research needs to better assess the actual impact. Large studies in homogeneous patient populations are required to address these problems.

## Data Availability

Data will be made available on publication and are available for review purposes on request from the authors.

## Declarations

### Ethics Approval and Consent to Participate

Not applicable

### Consent for Publication

Not applicable

### Availability of Data and Materials

The database on which this study is based is available on request from the authors.

### Authors’ Contributions

Both authors contributed equally. HW developed the protocol, ML edited and improved it and submitted it to PROSPERO. ML developed and implemented the search strategy. Both authors extracted the data independently, discussed discrepancies, and calculated the analyses independently. ML calculated the quantitative analysis reported and HW calculated the sensitivity analyses reported. ML wrote the first draft of the paper and HW edited and contributed to writing. Both authors analyzed and interpreted the data. All authors have read and approved the manuscript.

## Acknowledgment

Not applicable

## Author’s information

Harald Walach is a professor with Poznan Medical University, where he teaches mindfulness to the international medical students, and he is a visiting professor with University Witten-Herdecke, where he teaches philosophical foundations of psychology to psychology undergraduates. Apart from that he is founder and director of the Change Health Science Institute (www.chs-institute.org), based in Berlin Germany. Martin Loef is an independent researcher and Harald Walach’s partner in the CHS Institute, Berlin. He is a specialist in conducting systematic reviews and meta-analyses and lifestyle diagnostics.

